# An evaluation of the fracture resistance of endodontically treated extracted teeth restored with Titanium, Carbon and Fibre posts : An in-vitro randomized control trial

**DOI:** 10.1101/2020.04.13.20063883

**Authors:** Sonali Suvadarsini Behera, Gopal Krishna Choudhury, Abhilash Mohapatra, Sandeep Kumar Panigrahi

## Abstract

Post and core enforcement is done for Root Canal Treated (RCT) tooth as per need. Several posts and core systems are available claiming superiority. The usual post and core enforcement used in set up in India include that based on titanium and fiber. An unbiased in-vitro randomized control trial to compare the fracture resistance of RCT teeth with those restored with Titanium, Carbon and Fibre posts was hence conducted. The trial was conducted in a tertiary care dental institute in the eastern part of India. A total of 40 normal samples of extracted first mandibular were randomized to four groups (A through D) of ten samples each. Block random design was used, random number sequence was generated through computer centrally. The process was blinded, also during analysis. Group A (control) was RCT teeth with intact coronal structure while group B, C and D (interventional) were RCT teeth restored with titanium, carbon and fiber posts respectively. Universal Testing Machine (‘Tinius Olsen, H50KS’) was used for testing fracture resistance. The data were analyzed using Kruskal Wallis test followed by pairwise comparison in Stata v 12.1 SE. All statistical interpretations were done at p= 0.05 levels. The median fracture resistance among the groups was significantly different (p=0.000). Median fracture resistance of control, titanium, and fiber groups were similar (p >0.05). The median fracture resistance of the carbon group was inferior to all other groups (p<0.05). It was concluded that fiber and titanium post and core systems similar in strength to only root canal treated tooth, and scored better as compared to carbon post and core system.

## 1. INTRODUCTION

Definitive restoration in certain cases such as grossly decayed tooth, tooth having a short clinical crown, tooth with deep carries and trauma, etc. was not possible previously in the past. But with the advancement of material science, incurable tooth can now be restored to fully functional form using post and core system. These tooth are first endodontically treated and then followed by a post-core procedure which gives a predictable long term result.[1,2]

Root Canal Therapy (RCT) is known to be the treatment of choice for most of the decayed and damaged tooth. However, it gives a pulp-less tooth with less structural durability and fracture prone, owing to the cause of altered or decreased tooth structure for which RCT is carried out or due to instrumentation itself.[3,4] So, these teeth frequently require partial or complete-coverage restorations for esthetic and functional considerations. Type of restoration in a pulp-less tooth is determined by the amount of remaining tooth structure. Whenever the loss of clinical crown height is in horizontal pattern, it becomes mandatory to build up a complete core. Nevertheless, in such types of cases, a placement of post helps to maintain the core. Whenever there is loss of more than half of the coronal structures, post and core system are used for full coronal restoration. [5–7] Posts provide resistance and retention for core material whereas core provides stabilization to the corono-radicular part. The existence of a upright tooth structure in such case will provide a ferrule effect, and in turn will lead to long term accomplishment, weight sharing ability, improved steadiness and rotary motion confrontation.[2,8–10] The root is protected by the crown, which acts as a reinforcing ring or ferrule, to prevent vertical fracture of tooth, as has also been documented by artificial ferrule placement.[2,11]

The mean longevity of the post and core system as documented in studies is around 7.3 years. The cumulative rate of failure is around 11.3 years. Loss of retention is one of the most common causes of failure while posts having a high gold content seemed to have lower chances of failure compared to posts made of semi-precious alloy.[11–13] Researchers have shown that gold post and core is better with lower failure loads as compared to self-tapping dentin pins and either amalgam or composite as the core.[14] Some studies compared composite and amalgam direct cores sustained by an equivalent prefabricated pin and found both to be superior to a cast-gold core. [13,15] The procedure of using direct technique with prefabricated post and custom-made build-ups with composite resin or amalgam can also be completed in one appointment.

Several posts are available now-a-days with manufacturers of each of these posts claiming superiority. Studies have also been conducted which show varying results. Raju et al. found quartz fiber posts as having higher flexural strength compared to glass and composite-reinforced fiber posts. The higher flexural properties of fiber posts were also the reasons for prevention of root fracture.[7] Tortopidis et al. observed fiber-reinforced composite post were more esthetic.[16] Abduljawad et al. observed improved fracture resistance with glass fiber post in endodontically-treated teeth.[17] A study conducted by Maroulakos et.al. showed that titanium enforced post and core system and quartz fiber enforced post and core system were better than the gold standard cast gold post and core system.[18] However, there is no comparison of titanium, fiber and carbon post and core system.

There was insufficient literature comparing fracture-resistance of endodontically treated tooth restored with titanium, fiber and carbon as compared to the conventional root canal treated tooth, that was properly randomized for generating evidence.[19] The present in-vitro study was randomized in-vitro and designed to test the compressive resistance of endodontically treated extracted teeth restored with posts of titanium, carbon and fibre, as compared to controls that are not-treated with any post and known to have the poorest resistance.[20]

## 2. MATERIALS AND METHODS

The study was in-vitro, randomized control trial, conducted in the Department of Prosthodontics, Crown and Bridge and Implantology of a tertiary care dental institute of east part of India. Randomized control trial was perceived to provide progressive evidence as per the horizontal spectrum of evidence in prosthodontics.[21] The study was conducted from 2016-18, over a period of two years. Ethical permission for conducting the study was taken from the Institute Ethics Committee. Naturally extracted normal mandibular first pre-molar tooth which was extracted for orthodontic purpose was used for this study. Tooth of the participants belonging to the age group of 20 to 30 years, irrespective of gender, were used for this purpose to maintain homogeneity.

A total of 40 such teeth were taken for the study, based on sample size calculation in a similar study undertaken by Eapen et.al.[22] Root canal treatment was performed in all of these, and they were randomly allocated to four groups (A – D) based on previously generated random number sequence (block random sample design).[23] Group A consisted of only root canal treated tooth with intact coronal structure, group B consisted of titanium post and core, group C consisted of carbon post and core system while group D consisted of fiber post and core system. Control group was taken as RCT since it is generally the standard treatment.

Allocation to the groups was done by a person who was independent of the research, so as to prevent bias. The outcome assessment was similarly conducted by two independent observers, which further reduced observation bias. Considering the implications of following a protocol, we tried to decrease the lacunae and bias in an RCT conducted in-vitro settings using CRIS statement.[24,25]

### 2.1. Procedure for preparing the sample post RCT

#### 2.1.1. Group A

All teeth were mounted in acrylic resin at 130 degree angle up to Cemento-enamel junction(CEJ). Before mounting, approximately 0.25 mm thick light body was applied on the root portion to make it act as Periodontal ligament (PDL) fibers. Sealing of the access cavities was done using light cure composite resins.

#### 2.1.2. Group B

Sectioning of the crown was done by a fine grit diamond wheel perpendicular to the vertical axis of the tooth, in such a way that about 3 mm of coronal structure still remained from the CEJ. Then it was mounted in acrylic resin at 130 degree angle up to CEJ. Light body was applied on the root surface before mounting the teeth similar to the Group –A. Post space was prepared using Peeso-Reamer drill (available in titanium post kit) upto a maximum size of number 4. It was ensured to have a minimum of 4 mm of Gutta percha in the apical portion of the root. 37% phosphoric acid (META ETCHANT) was applied in the post space for a period of 15 seconds, and then rinsed off with the help of distilled water. Bonding agent (3M ESPE) was applied in the post space and then curing was done by using UV light for 10 seconds. Luxa Core Z (DMG) was injected into this canal and titanium post was placed into it by their individual post holder. Then 3 mm of core build up was done and cured using UV light. Then tooth preparation was carried out. A finish line of 0.5mm was given above the CEJ.

#### 2.1.3. Group C

Sectioning of the crown was done similar to the group-B and also mounted like that. Post space was prepared using Peeso-Reamer drill up to no-4 size. Rest of the procedure was same as that of group B. After injecting Luxa core Z (DMG) into the canal, carbon post was placed into the post space. Core build up and finish line was done as in group B.

#### 2.1.4. Group D

Fiber post was placed into the post space after post-space preparation. Rest of the procedure were similar as in group B and C.

### 2.2. Procedure for testing samples for fracture resistance

A computer controlled Universal Testing Machine (“Tinius Olsen, H50KS”) was used for testing fracture resistance. Specimens along with acrylic block were mounted using a special fixture on these testing machines. A compressive load was applied at 130 degrees to the vertical axis of the tooth to evaluate the load bearing capacity using ball-ended steel compressive head having a diameter 0.5 mm/min. The resistance capacity was noted at the point when the sample gave way.

### 2.3. Data analysis

Fracture resistance was expressed in Kilo Newtons (KN). Data were entered in an excel sheet (Microsoft Excel v. 2016), and imported to licensed version of Stata 12.1 SE for analysis. Considering the small sample size in each group (n=10), quantitative data were expressed as median with inter-quartile range. Median fracture resistance of the groups was analyzed using Kruskal-Wallis test (non-parametric test) followed by pairwise comparison of medians between the groups to find out if there was any significant difference between any of the groups which was finally leading to a significant difference. All statistical interpretations were done at α = 0.05 levels.

## 3. RESULTS

The median fracture resistance was highest for fiber (0.776 KN) and lowest for carbon (0.482 KN) and followed the following order (fig 1): **Fiber (Gr D) >> Titanium (Gr B) >> Control (Gr A) >> Carbon (Gr C)**.

**Fig. 1:**
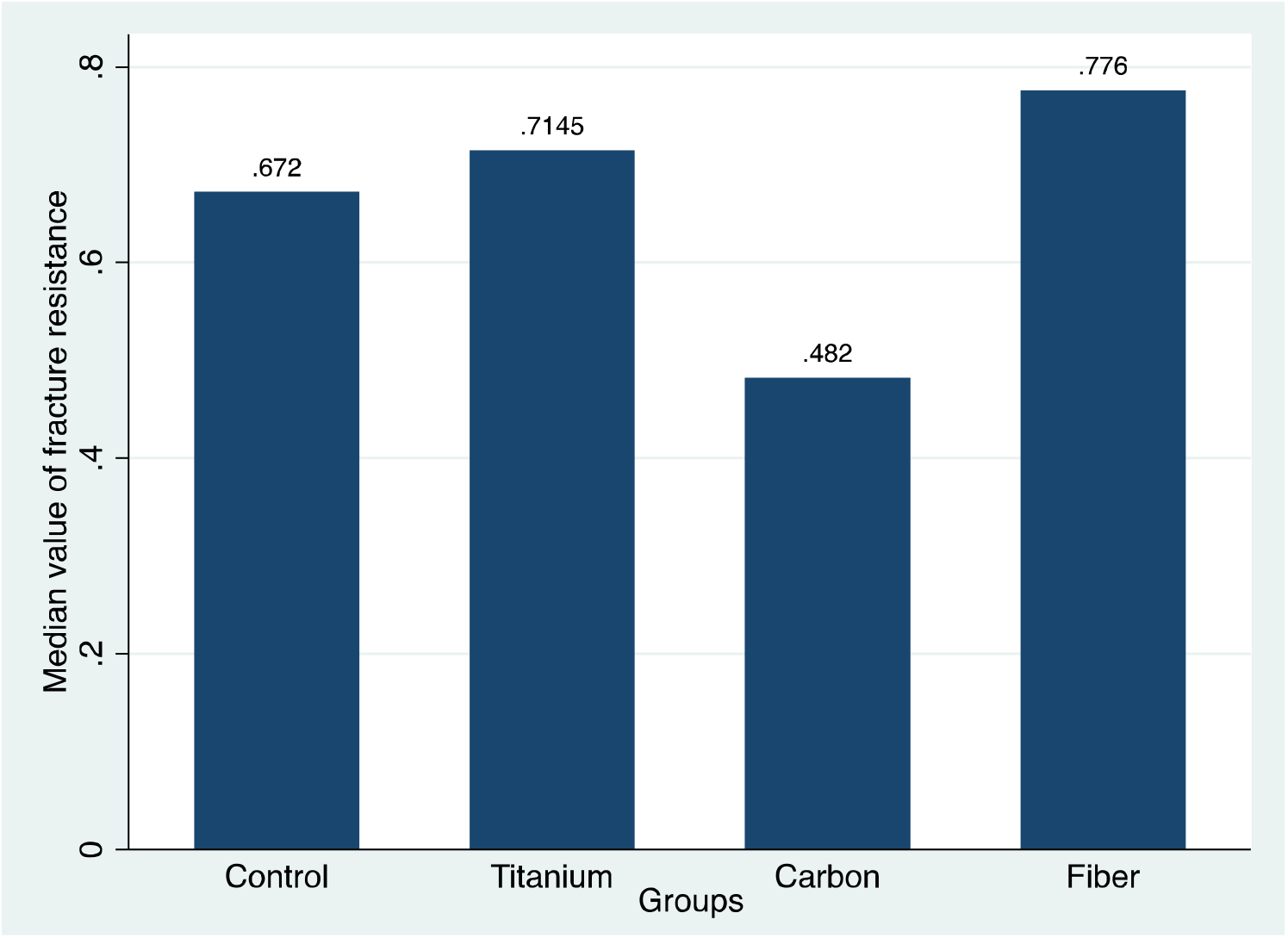
Comparison of median values of the fracture resistance among the groups (n=10 each)

The median fracture resistance (in KN) of the groups were compared using Independent Sample Kruskal-Wallis test which showed that a statistically significant difference was present (p=0.000, Kruskal Wallis test) (table 1, and fig 2).

**Table 1:** Comparison of median fracture resistance of the study groups (n=10 each)

| Sl | Group | Median fracture resistance (in KN) | IQR (Q1, Q3) | P value |
| --- | --- | --- | --- | --- |
| 1 | Control (Gr A) | 0.672 | 0.601, 0.793 | 0.000 |
| 2 | Titanium(Gr B) | 0.714 | 0.683, 0.789 |  |
| 3 | Carbon(Gr C) | 0.482 | 0.452, 0.554 |  |
| 4 | Fiber (Gr D) | 0.776 | 0.697, 0.875 |  |

**Fig 2:**
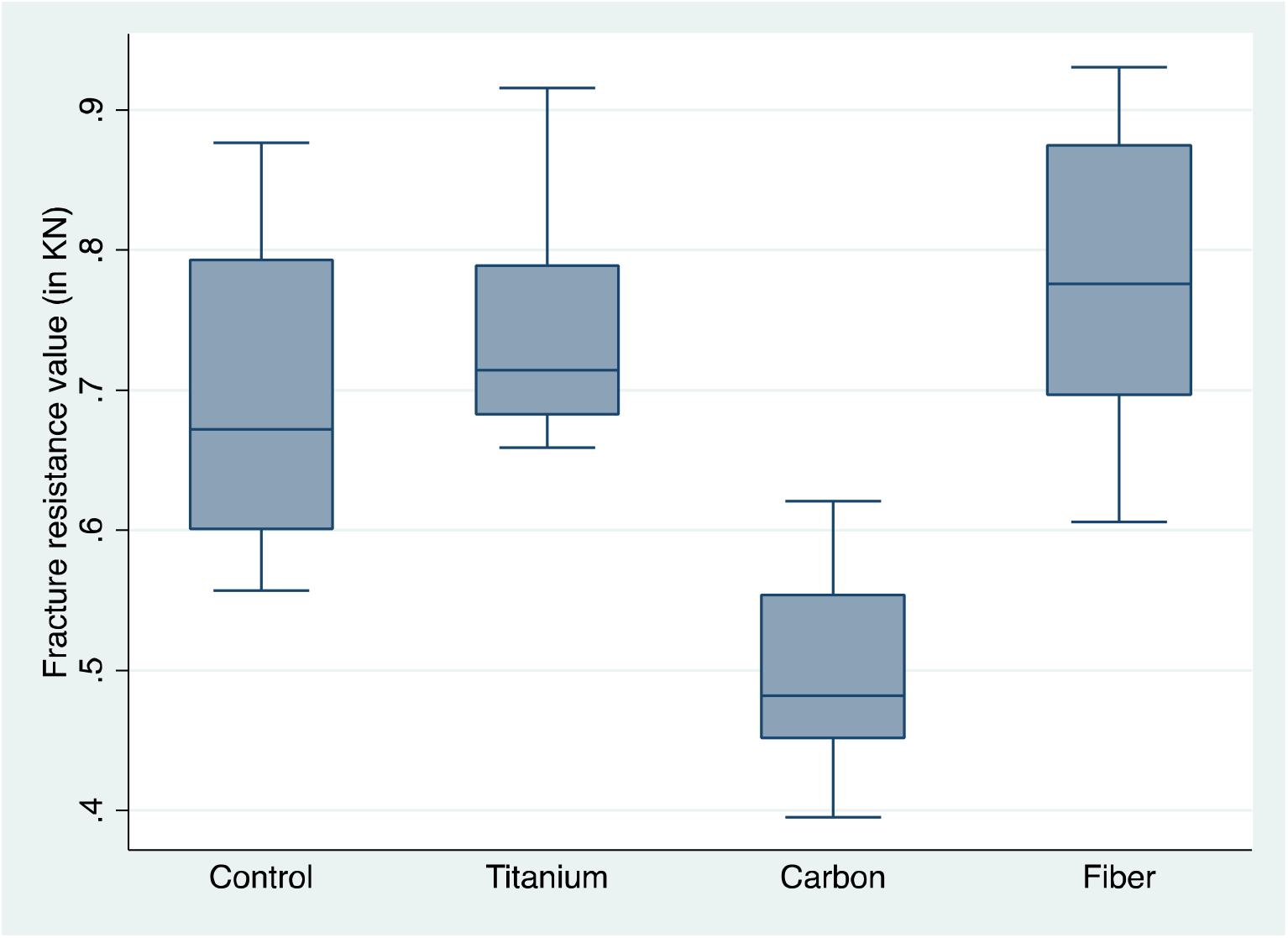
Comparison of median values of fracture resistance in between the groups (n=10 each)

Differences in between which groups might have contributed to the significant difference in the whole group was tested using pairwise comparisons, and it was seen that carbon group was quite inferior to all other groups, and there was a significant difference between Carbon – Control, Carbon – Titanium and Carbon – Fiber (p<0.05). However, Control group, Titanium group and Fiber group were all comparable as can be seen in table linked to Fig 2 and table 2 (where p>0.05).

**Table 2:**
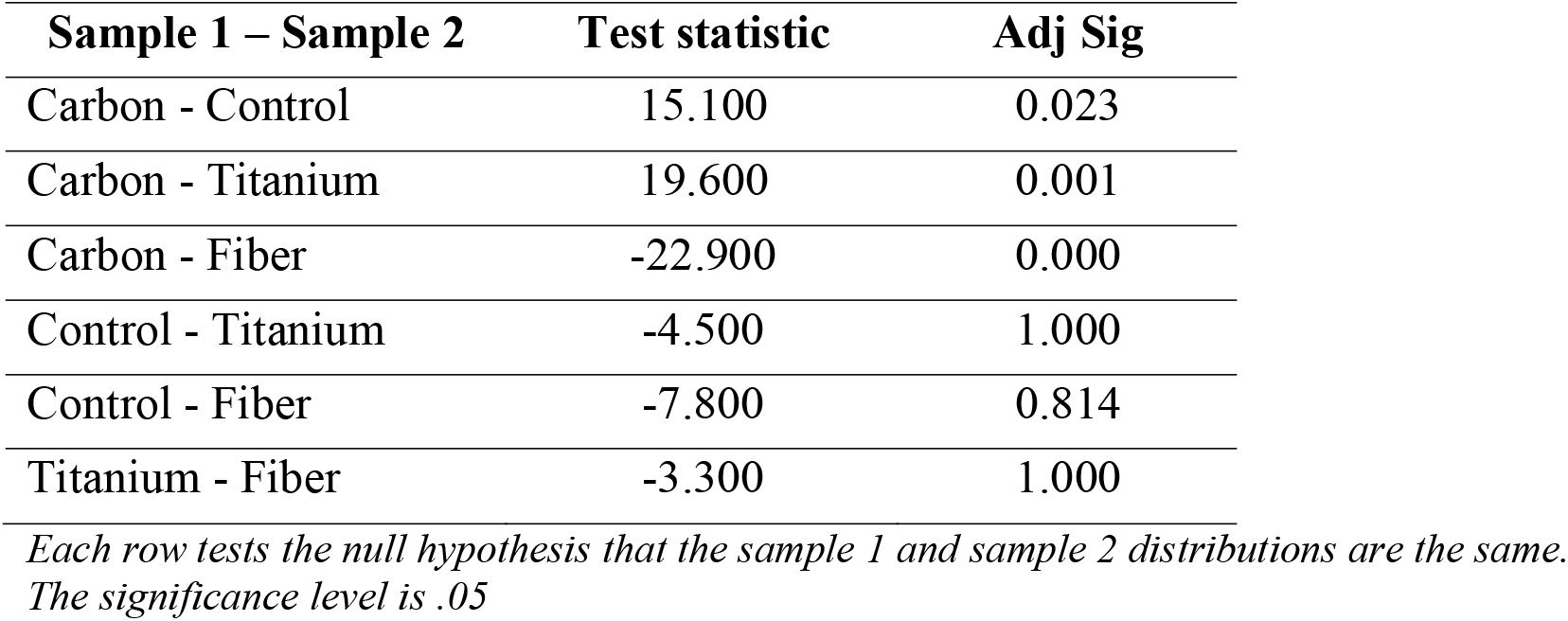
Pairwise comparisons in between the groups (n=10 each)

Thus the final order of the differences of fracture resistance of the models as evident from the fig. 2 was found to be:

**Fiber (Gr D) == Titanium (Gr B) == Control (Gr A) >> Carbon (Gr C)**

## 4. DISCUSSION

The way the tooth structure is extended coronally above the crown margin decides failure threshold to the most. A minimum of 1.5 mm ferrule height is sufficient enough to ensure restoration prognosis in a favourable way, and presence or absence of ferrule is the main factor determining the success rate of any post and core with a success rate ranging from 0% to 97% for posts without a ferrule. Similarly, the survival of crown without posts is usually in a range of 0% to 100%.[26] In this study, post and core were used along with a ferrule of 3 mm thickness to ensure a consistent success/ survival rate. As it was an in vitro study the the survival rate could not be tested, though ensured ethically.

Fracture resistance was tested using a Universal testing machine subjecting to a load of 0.5 mm/min at an angle of 130 degrees. An angle of 130 degrees is considered for testing strength of the samples since majority of the fractures occur due to angular force in reality. Control group selected were samples of 10 endodontically treated mandibular first premolar. It was because of the reason that RCT currently is the most common and standard procedure adopted for crown placement.

In this study, the fracture resistance of teeth that were restored with three different post systems were compared. Teeth extracted for orthodontic treatment were used for the preparation of the specimens. All the teeth received the endodontic treatment, and standard cores (paracore) were fabricated. A layer of silicon layer approximately 0.25mm (light body) to simulate periodontal ligament was created for limited freedom of movement. Then they were stored in normal saline for 30 days to simulate the oral cavity environment. Resin cement was used for these posts for silanating and bonding, as in few other studies.[27] Amalgam vs composite core build up have shown that composite core build up fares better result. So core build up material Paracore (Densply, USA) was used in this study.

The comparison of median fracture resistance between experimental groups were carried out and statistical analysis done. A significant difference in fracture resistance was seen among the groups tested (p=0.000, Kruskal Wallis test). Pair wise comparison showed that the significant difference was due to the difference in between Carbon-Control, Carbon-Titanium and Carbon-Fiber pairs. However, Control group, Titanium group and Fiber group were all comparable (where p >0.05). Thus the final order of the differences of fracture resistance was: **Fiber (Gr D) == Titanium (Gr B) == Control (Gr A) >> Carbon (Gr C)**.

Metal alloys used to form all the pre-fabricated posts, leading to a final mix of materials made up of dentin, cement and metallic post and core. These systems had a major disadvantage of having uncontrolled stress at the root.[28] Dentistry now uses more of metal-free homogenous materials while restoring endodontically treated tooth, whose physical properties are like that of dentin. One of such post systems is that reinforced by fiber. The modulus of elasticity of several metallic posts have been found to be more than 20 times (around 220,000 MPa) that of dentine, whereas that of glass fiber posts (54000 MPa) are more nearer to that of dentine (20000 MPa), thus reducing the chances of root fracture.[5,6,29] Glass-fiber post systems are made up of glass fibers in a single direction within the resin matrix, thus enhancing the strength of the post without a change in the modulus of elasticity.[27]

Few studies have demonstrated carbon post and core as having better strength and bondability.[5,6,29] But in the present study it was not found to have a high compressive strength in comparison to other groups. However, a study conducted by Sharma et.al, however had earlier demonstrated that quartz fiber treated tooth had better resistance compared to carbon treated ones, similar to the present study.[20]

Posts like fiber having high elastic modulus had a high range of survival/ success (between 71.8% to 100%), [26] and fiber showed high modulus.[30] Thus, evidence says that fiber should have a good success/ survival when used as post and core. Titanium being one of the strongest metals do not bend under stress and therefore if fractures, fractures the roots.[18] Glass fibre is a material which has a better modulus of elasticity therefore is more resistant to fracture and is as good as a endodontically treated teeth. [7] In this study setting, however, the results were found to contradict other studies. Fiber post and core was not found to be similar to titanium post and core, or normal RCT tooth.

The fracture resistance test was limited to in vitro results, and sometimes it may not corroborate with the clinical results in vivo. This may be due to reasons such as environmental factors that may interact and affect the performance of a dental implant, eg. bacterial flora, especially late colonizers doing more damage to dental implants than early ones.[31] There is not much difference in the dental materials, however, considering response of these oral pathogens growing on implants to antiseptics.[32] Appropriate in-vivo experimental study need to be done to arrive at clinical conclusion.

## 5. CONCLUSION

The study concluded that there was the significant difference in the fracture resistance between the post and core of fiber, titanium, carbon and control groups (p<0.05). This was basically due to the differences between carbon post and core and other three groups (titanium, fiber and control) (p<0.05). The fracture resistances were almost similar between control (RCT tooth with intact coronal structure), fiber and titanium groups of post and core (p>0.05). Carbon, was thus the least desirable material to be used for post core build up among all, while fibre posts was the best. Further in vivo studies in line with the study may be needed to enforce the findings in clinical practice.

## Data Availability

Data is available with the corresponding author and can be produced for personal use on request. Please send requests for data, quoting intent of use and acknowledgment to

## Acknowledgements

Nil

## Ethical statement

Ethical clearance was taken from the Institute Ethics Committee of IMS and SUM Hospital, Siksha ‘O’ Anusandhan deemed to be University. Informed consent was taken from the patients to use the removed tooth for in-vitro studies. In the total process no harm was done to any human or animal subject.

